# Burden, Risk Factors, and Clinical Outcomes of Pediatric Malaria in Nigeria: A Systematic Review and Meta-Analysis Protocol

**DOI:** 10.1101/2025.10.22.25338582

**Authors:** Olayinka Rasheed Ibrahim, Jubril Abdulkareem, Amudalat Issa, Michael Abel Alao

## Abstract

**Background:** Nigeria ranks number one globally in malaria burden, with the exact burden, especially for hospitalization, unknown. This systematic review and meta-analysis intend to fill this gap by pooling the analysis of pediatric malaria and associated factors in Nigeria.

**Methods and Analysis:** A comprehensive search strategy using MeSH terms, text words, and entry terms will be applied to six databases: PubMed, Embase, EBSCO Hosts, Google Scholar, Web of Science, and Scopus. Eligible studies will be observational and published in English from inception till June 30, 2025. The primary outcome is the pooled prevalence of pediatric malaria [overall, severe malaria vs. uncomplicated]. Secondary outcomes include factors influencing clinical presentation, severity, and outcomes, as well as the effects of moderators such as age, sex, socioeconomic status, and geographic location. Data extraction will capture study characteristics, participant demographics, and outcome measures. Methodological, clinical, and statistical heterogeneity will be assessed. Risk of bias will be evaluated using the Newcastle-Ottawa Scale. Publication bias will be examined using funnel plots, Egger’s regression. Pooled estimates will be reported with 95% confidence intervals. This will summarize the data on pediatric malaria in Nigeria. Using the random-effects models, the pooled prevalence along with the 95% CI and the *I*^2^ for the test of heterogeneity will be reported. We will report the meta-regression analysis of factors influencing pediatric malaria in Nigeria.

**Conclusions:** This study will provide robust data on pediatric malaria and associated factors in Nigeria. The findings from this study will inform the country’s policy and public health approach as the nation strives to eliminate malaria in line with the WHO’s goals.

**PROSPERO Registration Number:** CRD420251163322

## Introduction

Malaria is a parasitic infection caused by *Plasmodium spp*., which has remained a disease of public health significance in the tropics. The 2024 World Health Organization (WHO) malaria report estimated 263 million (95% confidence interval 238 million to 294 million) malaria cases and 597,000 related deaths in 2023.[1] The significant burden of malaria disproportionately affects sub-Saharan Africa, with Nigeria ranked as number one and accounting for 27% of the global cases.[2] In 2023, the country reported an estimated 68.1 million (95% confidence interval: 49.1 million to 92.6 million) malaria cases and approximately 185,000 deaths.[1] In Nigeria, like the rest of the world, the significant burden of malaria affects the pediatric age group, where it is as high as 80%.[2]

Despite the available estimates on malaria in Nigeria, the data do not delineate to reflect the actual burden of pediatric severe malaria at the hospital levels, uncomplicated malaria, associated factors, various complications, and clinical outcomes.[1, 2] Available studies conducted among children and adolescents in Nigeria have demonstrated significant variations in the local burden, complications, and outcome of pediatric malaria across the six geopolitical zones in Nigeria and within the same zone. [2–5] With variations in the burden reported across the country, the exact burden of severe malaria, uncomplicated malaria, associated factors, complications, and clinical outcomes in the country remains largely unknown, which hampers resource allocation and policy formation.

This observation, therefore, brought forward the need to synthesize the data on the various studies conducted on the Nigerian pediatric population with malaria and associated factors. The synthesis of this data will also enable the examination of trends over the years, assess the country’s progress, provide information for policy interventions, and ultimately ensure that the country achieves its goal of eliminating malaria. We therefore raised the following research questions: What is the pooled prevalence of pediatric malaria in Nigeria [severe and uncomplicated]? What are the associated factors for pediatric malaria in Nigeria? What is the trend of pediatric malaria in Nigeria [severe and uncomplicated]? What are the outcomes [discharged, deaths, and complications, e.g., neurological] of pediatric severe malaria in Nigeria?

This study intends to conduct a systematic review and meta-analysis of studies on pediatric malaria [severe and uncomplicated] in Nigeria, trends, associated factors, and clinical outcomes [discharge, deaths, and complications] in Nigeria.

## Methods

The review was registered with the International Prospective Register of Systematic Reviews (PROSPERO: CRD420251163322) to enhance methodological transparency and avoid duplication of research. Any protocol amendments will be documented and updated accordingly in the registry.

### Eligibility criteria

The PICOS will be used to define the eligibility criteria for this study and will include:

Population (P) —Published studies conducted in Nigeria that include children and adolescents aged 0 to 18 years diagnosed with malaria (severe and uncomplicated malaria)

Intervention (I) —Not applicable based on the topic outline. However, exposures such as socioeconomic factors, age, sex, geopolitical zones, and diagnostic methods will be examined as potential associated factors.

Comparator (C) —Comparisons across sex, age groups, socioeconomic class, geopolitical zones, diagnostic methods, and Plasmodium species.

Outcomes (O) —Primary outcome will include

1. The pooled prevalence of pediatric malaria [overall, severe malaria vs. uncomplicated].

The secondary outcomes

1. The associated factors for pediatric malaria
2. Outcomes of severe malaria, including discharge, mortality, and complications (e.g., neurological sequelae)
3. The trends of pediatric malaria over time
4. Subgroup analysis of prevalence by age, sex, socioeconomic status, and geopolitical zones.

Study design (S): This review will include quantitative observational and interventional studies, including cross-sectional, cohort, case-control, and longitudinal studies, conducted in Nigeria and published in any language. Non-English studies will be translated using professional translation tools, Google Translate (academic mode) and Microsoft Translator. The translated studies will be verified by native-language speaker to ensure accuracy during data extraction

### Exclusion criteria

The following studies will be excluded from this study

1. Case reports, case series, editorials, commentaries, qualitative studies, and review articles (including narrative and systematic reviews)
2. Studies without clearly defined population denominators
3. Studies that are irretrievable after reasonable attempts to contact the corresponding authors
4. Studies with a sample size of fewer than 100 participants
5. Studies without laboratory confirmation of malaria [microscopy, rapid diagnostic tests for malaria, or polymerase chain reaction]
6. Studies on asymptomatic malaria

### Information sources

A comprehensive literature search will be conducted across the following electronic databases: PubMed, Embase, EBSCO Hosts, Google Scholar, Web of Science, and Scopus. In addition, manual searches will be performed by screening the reference lists of studies that will be included and relevant reviews to identify any additional eligible articles and grey literature sources.

### Search strategy

The search strategy will combine Medical Subject Headings [MeSH] terms, keywords, and entry terms adapted for each database to ensure sensitivity and specificity. The terms and keywords will include variations and combinations of the following: Geographic terms: “Nigeria”; Population terms: “newborns,” “infants,” “children,” “adolescents”; Disease terms: “malaria,” “severe malaria,” “uncomplicated malaria”; Outcome and associated factor terms: “sex,” “socioeconomic status,” “geopolitical zones,” “associated factors,” “complications,” “outcomes,” “discharge,” “deaths,” and “hospitalization.” An example of the PubMed search string (to be adapted for other databases) is as follows: (“Nigeria”) AND (“children” OR “infants” OR “newborns” OR “adolescents”) AND (“malaria” OR “severe malaria” OR “uncomplicated malaria”) AND (“complications” OR “outcomes” OR “associated factors” OR “socioeconomic status” OR “sex” OR “geopolitical zones” OR “discharge” OR “death” OR “hospitalization” OR “outcomes”). The search period will be inception till June 30^th^, 2025.

There will be no language restrictions during the literature search. All relevant studies, irrespective of language, will be included. Non-English studies identified during the screening will be translated to English before data extraction to minimize selection and translation bias Study

### Selection

Following the literature search, all retrieved records will be exported into the Rayyan software for systematic screening and management. The screening will be done using different levels to enhance transparency as follows: Level 1 would involve screening of identified studies for the study design quantitative observational and interventional studies would be accepted; Level 2 will involve screening of identified studies in the titles and abstracts using entry terms, keywords and MeSH terms; Level 3 will involve further screening of the contents of articles by reading the full article using the same search strategy; Level 4 will involve snowballing of literature on references from included studies; Level 5: Studies will be screened at outcome levels to select those that reported the primary outcome with or without secondary outcomes; Level 6 will involve grey literature that report primary outcome and or secondary outcomes. The screened will be done independently by two reviewers (AI and JA) to identify studies that potentially meet the inclusion criteria. Any discrepancies or disagreements between the two reviewers will be resolved by a third reviewer (ORI). The study selection process will adhere to the Preferred Reporting Items for Systematic Reviews and Meta-Analyses (PRISMA 2020) guidelines.

### Data collection process

Data extraction will be carried out independently by two reviewers [AI and JA] using a predefined data extraction form that will be created in Microsoft Excel. The data will include the first author’s name, year of publication, total number of study participants, number of cases [severe/uncomplicated malaria], study settings, study design, diagnostic methods for malaria, age, sex, clinical outcomes (e.g., discharge, death, neurological sequelae), states/geopolitical zones of the studies, associated factors (demographics, socioeconomic status, environmental and climatic factors (rainfall pattern/seasonality, rural vs. urban), ownership of bed nets, utilization of bed nets, and parasite species). Any discrepancies in the extracted data will be resolved in consultation with the fourth reviewer (MAA).

#### Operational definition

The definitions of uncomplicated malaria and severe malaria will follow the WHO definitions.[6] Uncomplicated malaria is defined as the presence of malaria symptoms along with confirmed malaria parasites, but without any signs of vital organ dysfunction.

Severe malaria: Presence of *Plasmodium falciparum* parasitemia with vital organ dysfunctions, as indicated in the WHO guidelines for severe malaria

### Risk of bias assessment and methodological quality assessment

The methodological quality and risk of bias of the included studies will be independently assessed by two reviewers [AI and JA] using the Newcastle-Ottawa Scale (NOS) adapted for observational studies, while Cochrane Risk of Bias tool (RoB 2.0) will be used for interventional studies.[7] The NOS tool evaluates observational studies based on three key domains: selection of study groups, comparability, and ascertainment of outcomes or exposures. Each study will be assigned a score based on the NOS criteria, and the results will be categorized as low, moderate, or high risk of bias. Inter-rater agreement between the two reviewers will be assessed using Cohen’s kappa statistic to assess the consistency of screening and risk of bias assessment. Where significant differences exist between the two rates, the third reviewer (ORI) will serve as an arbiter, and their final rating will be used for the study.

### Data Synthesis and Statistical Analysis

Data extracted using the predefined Excel spreadsheet will be imported into R statistical software for analysis, using the “meta” and “metafor” packages. The pooled prevalence of pediatric malaria (overall, severe, and uncomplicated) will be estimated, along with 95% confidence intervals (CIs). The odds ratio (OR) along with 95% CI will be used to express factors that are associated malaria. If significant heterogeneity is observed among the included studies, a random-effects model (DerSimonian–Laird method) will be applied to compute the pooled prevalence estimates, providing a more conservative measure of variability. Forest plots will be generated to visually display the pooled estimates, corresponding confidence intervals, and prediction intervals.

### Assessment of Heterogeneity

Statistical heterogeneity among studies will be evaluated using the Cochrane’s Q test and quantified with the *I*^2^ statistic.[8] An *I*^2^ value greater than 50% will be considered indicative of substantial heterogeneity. Where substantial heterogeneity exists, subgroup analyses and mixed-effect meta-regression will be conducted to explore potential sources of variation, such as age group, sex, socioeconomic status, geopolitical zone, diagnostic methods, and study design.

### Assessment of Publication Bias and Sensitivity Analysis

Publication bias will be evaluated visually using funnel plots and Egger’s test. To assess the potential influence of missing studies, a trim and Fill analysis will be performed. Sensitivity analyses will be conducted to evaluate the impact of studies with higher risk of bias on the pooled estimates. Additionally, a leave-one-out sensitivity analysis will be performed to assess the influence of each study on the overall pooled estimate and to evaluate the robustness of the findings.

All statistical tests will be two-tailed, and a p-value < 0.05 will be considered statistically significant for all analyses.

### Status and timeline of the study

This protocol has just been registered at PROSPERO with preliminary search of title related to the work to avoid duplication of previous work. Afterward, the plan is to publish the protocol, conduct literature search, data abstraction, analysis and sharing the findings of the study [Table 1].

**Table 1:** Propose timeline for the systematic review and meta-analysis.

| Month | September<br>2025 | October<br>2025 | November<br>2025 | December<br>2025 | January<br>2026 | February<br>2026 |
| --- | --- | --- | --- | --- | --- | --- |
|  | Title search<br>and<br>registration<br>(done) | Submission<br>for peer<br>review<br>(done) | Literature<br>search (yet to be<br>done) | Data<br>abstraction<br>(yet to be<br>done) | Data<br>analysis<br>(yet to be<br>done) | Submission<br>of finding<br>for<br>publication<br>(yet to be<br>done) |

### Ethical consideration and declaration

As this study involves the review and synthesis of data from previously published studies, ethical approval is not required. No primary data will be collected, and the review will not involve any direct interaction with human participants or access to confidential patient information

## Results

The results of this systematic review and meta-analysis will be reported in accordance with the Preferred Reporting Items for Systematic Reviews and Meta-Analyses (PRISMA) 2020 guidelines. The findings from this study will be a detailed description of the literature search process, the number of records retrieved from each database, the total number of studies screened, and the number of studies included in the final analysis. A table summary of characteristics of the final studies included in the analysis will capture the number of participants, study design, malaria classification, geographic distribution, associated factors, diagnostic methods, complications, and clinical outcomes (e.g., discharge, death, neurological sequelae) and risk of bias of each study.

For the data synthesis, the random-effects model will be used to estimate the pooled prevalence and 95% confidence intervals (CIs) for overall malaria, severe malaria, uncomplicated malaria, complications of severe malaria, associated factors, and clinical outcomes.

Subgroup analysis will include the burden in various age groups, sex, socioeconomic status, geographical locations, seasonality, use of insecticide-treated nets, study settings, study design, diagnosis, and year of publication.

A meta-regression analysis will be employed to assess the influence of these subgroup variables and other study-level covariates on the pooled prevalence estimates, thereby identifying factors that may contribute to heterogeneity across studies.

The findings will be displayed as figures (forest plots, funnel plots, fill and trim plots) and summary tables.

## Discussion

The findings from this systematic review and meta-analysis will provide a comprehensive and evidence-based estimate of the actual burden of pediatric malaria in Nigeria, encompassing both severe and uncomplicated forms. By synthesizing data from multiple studies across diverse settings and populations, this review will fill a critical knowledge gap in the epidemiology of malaria in pediatric age groups in Nigeria.

The results will be contextualized and compared with existing regional and global malaria data, highlighting similarities and differences in prevalence, associated factors, and outcomes. Potential explanations for observed variations—such as differences in diagnostic methods, transmission intensity, socioeconomic factors, and healthcare access—will be explored to provide a comprehensive overview of malaria patterns in Nigeria’s pediatric population.

The findings from the meta-regression analysis are expected to identify and model key determinants of malaria prevalence and severity, such as age, sex, socioeconomic status, geographic zone, and diagnostic method. These insights will be invaluable in informing policy formulation and revision, particularly as Nigeria continues to align its malaria control and elimination strategies with the WHO’s global malaria targets.

The research, policy, and public health implications of this study will also be discussed in detail. Specifically, the study is expected to guide resource allocation, targeted interventions, and surveillance strategies for malaria prevention and management in children.

Finally, the discussion will also address the limitations of the review, including potential publication bias, heterogeneity in study methodologies, and regional data gaps, to ensure a transparent interpretation of the findings.

## Dissemination

The findings from this systematic review and meta-analysis will be disseminated widely to maximize their impact. Results will be submitted for publication in a peer-reviewed scientific journal and presented at national and international conferences related to infectious diseases, pediatrics, and public health. In addition, summaries of the findings may be shared with relevant stakeholders, including policymakers, malaria control programs, and public health institutions, to support evidence-based decision-making and malaria elimination efforts in Nigeria. All data supporting the findings of this review will be made available in the published article and its supplementary materials, ensuring transparency and reproducibility.

## Data Availability

The data that will be available for this study, will deposit in public repository

## List of Abbreviations

PRISMA-P: (Preferred Reporting Items for Systematic review and Meta-Analysis Protocols)

NOS: Newcastle–Ottawa Scale
PROSPERO: International Prospective Register of Systematic Reviews

## Declarations

### Authors’ contributions

ORI conceived the study, was involved in the study design, literature review, search, data extraction, analysis, draft of the manuscript and approved the protocol.

JA was involved in the study design, literature review, search, data extraction, analysis, draft of the manuscript and approved the protocol.

AI was involved in the study design, literature review, search, data extraction, analysis, draft of the manuscript and approved the protocol

MAA conceived the study, was involved in the study design, literature review, search, data extraction, analysis, draft of the manuscript and approved the protocol

## Acknowledgements

We will acknowledge any individual that help in this work but does not fulfil authorship criteria as listed by International Committee of Medical Journal Editors.

## Support information

PRISMA-P checklist-uploaded

Search strategy-uploaded

a. The review work is self-funded

b.Sponsor: All the authors contributed to sponsor the project

c.Guarantor of the review: ORI

